# Risk of death among people with rare autoimmune diseases compared to the general population in England during the 2020 COVID-19 pandemic

**DOI:** 10.1101/2020.10.09.20210237

**Authors:** Emily Peach, Megan Rutter, Peter Lanyon, Matthew J Grainge, Richard Hubbard, Jeanette Aston, Mary Bythell, Sarah Stevens, Fiona Pearce

**Affiliations:** Division of Epidemiology and Public Health, University of Nottingham UK; Nottingham University Hospitals NHS Trust, Nottingham, UK; National Disease Registration Service, Public Health England

**Keywords:** COVID-19, Rare autoimmune rheumatic diseases, Lupus, Vasculitis, Scleroderma, Myositis, Juvenile idiopathic arthritis, Epidemiology, Mortality

## Abstract

**Objectives:** To quantify the risk of death among people with rare autoimmune rheumatic diseases (RAIRD) during the UK 2020 COVID-19 pandemic compared to the general population, and compared to their pre-COVID risk.

**Methods:** We conducted a cohort study in Hospital Episode Statistics for England 2003 onwards, and linked data from the NHS Personal Demographics Service. We used ONS published data for general population mortality rates.

**Results:** We included 168,691 people with a recorded diagnosis of RAIRD alive on 01/03/2020. Their median age was 61.7 (IQR 41.5-75.4) years, and 118,379 (70.2%) were female. Our case ascertainment methods had a positive predictive value of 85%. 1,815 (1.1%) participants died during March and April 2020. The age-standardised mortality rate (ASMR) among people with RAIRD (3669.3, 95% CI 3500.4-3838.1 per 100,000 person-years) was 1.44 (95% CI 1.42-1.45) times higher than the average ASMR during the same months of the previous 5 years, whereas in the general population of England it was 1.38 times higher. Age-specific mortality rates in people with RAIRD compared to the pre-COVID rates were higher from the age of 35 upwards, whereas in the general population the increased risk began from age 55 upwards. Women had a greater increase in mortality rates during COVID-19 compared to men.

**Conclusion:** The risk of all-cause death is more prominently raised during COVID-19 among people with RAIRD than among the general population. We urgently need to quantify how much risk is due to COVID-19 infection and how much is due to disruption to healthcare services.

**Key messages:**

- People with RAIRD had an increased risk of dying during COVID-19 from age 35 years onwards, whereas in the general population it increased from the age of 55 onwards.
- Women had a greater increase in their risk of death during COVID-19 compared to men.
- The risk of working age people with RAIRD dying during COVID-19 was similar to that of someone 20 years older in the general population.

## Introduction

There are limited data, and no whole population studies, on the impact of COVID 19 on people with rare diseases. This is important to inform shielding advice for people with rare diseases, and also to communicate realistic levels of risk as they apply to different groups and allow informed personal choices (1). There is also a risk that the known discrepancy in outcomes and access to treatment between rare and common diseases will widen, leading to more health inequality(2).

We have studied the risk of death during the 2020 UK COVID-19 epidemic across rare autoimmune rheumatic diseases (RAIRD) because they are an exemplar group of rare diseases requiring treatment with immunosuppression. People with these conditions have slightly increased mortality compared to the general population, and they may be at increased risk both due to COVID-19 infection and from their disease when healthcare services are disrupted. However, their risk during COVID-19 has not been quantified. Using linked national health records for the whole population of England we have estimated the risk of death among people with rare autoimmune rheumatic diseases during March and April 2020, at the beginning of the COVID-19 pandemic in the UK, and compared this to 1) their risk before COVID-19 and 2) the risk of death in the general population during COVID-19.

## Methods

The Registration of Complex Rare Diseases Exemplars in Rheumatology (RECORDER) project is a collaboration between the University of Nottingham, and the National Congenital Anomaly and Rare Disease Registration Service (NCARDRS) within Public Health England. NCARDRS registers people with congenital abnormalities and rare diseases across the whole of England. Currently there is no routine notification to NCARDRS by healthcare providers of people with rare diseases that are non-genetic and occur after infancy(3,4). RECORDER has established the methodologies for identification, validation and registration with NCARDRS of people who have later onset, non-genetic rare diseases, using rare autoimmune rheumatic diseases as an exemplar group of conditions. This has been enabled through NCARDRS access to national routinely collected healthcare datasets, such as Hospital Episode Statistics (HES). HES contains every episode of admitted NHS patient care in England (in-patient and day-case), with all prevalent diagnoses coded according to ICD-10. Importantly, each of the rare autoimmune rheumatic diseases maps to a unique ICD-10 code that does not also include other conditions, and patients with these conditions also have frequent in-patient or day-case activity, making them ideal for identification in HES.

We have previously validated ascertaining diagnoses of vasculitis (ANCA-associated vasculitis, Takayasu arteritis and Kawasaki disease) in HES, with positive predictive values (PPV) over 85%(5). For this study we have used NCARDRS legal permissions and data sharing agreements with NHS Trusts in England to validate additional diagnoses, confirming diagnoses of randomly selected people with coded diagnoses of systemic lupus erythematosus, scleroderma, idiopathic inflammatory myositis, Behcet’s disease, giant cell arteritis and juvenile idiopathic arthritis in two hospital Trusts.

We included people who had a diagnostic code for a rare autoimmune rheumatic disease in in-patient HES from 2003 onwards, were resident in England, and who were alive on 1 March 2020. We used data from the NHS Personal Demographics Service, linked by NHS number and date of birth, to ascertain whether people were alive or dead, and their date of death(6). Authors extracted data from the whole HES dataset themselves. A data flow diagram is shown in figure 1. The data did not require further cleaning after building the cohort. We calculated the mortality rate during March and April 2020, using the cohort of people with rare autoimmune rheumatic conditions as the denominator population. We repeated the calculation for mortality rates during March and April for each of the 5 previous years for comparison. We converted all rates into age-standardised mortality rates (ASMRs) per 100,000 population, standardised to the 2013 European Standard Population. We calculated sex-specific mortality rates, including ASMRs however it should be noted that the European standard population is not disaggregated by sex, meaning it assumes equal numbers of males and females, and identical distributions by age for males and for females. The World Health Organisation and the Office for National Statistics (ONS) therefore advise focusing on age-standardised rates rather than age-sex standardised rates for headline measures. Age-specific mortality rates per 100,000 people were calculated in 10-year age bands for comparison to data on the whole population of England.

**Figure 1:**
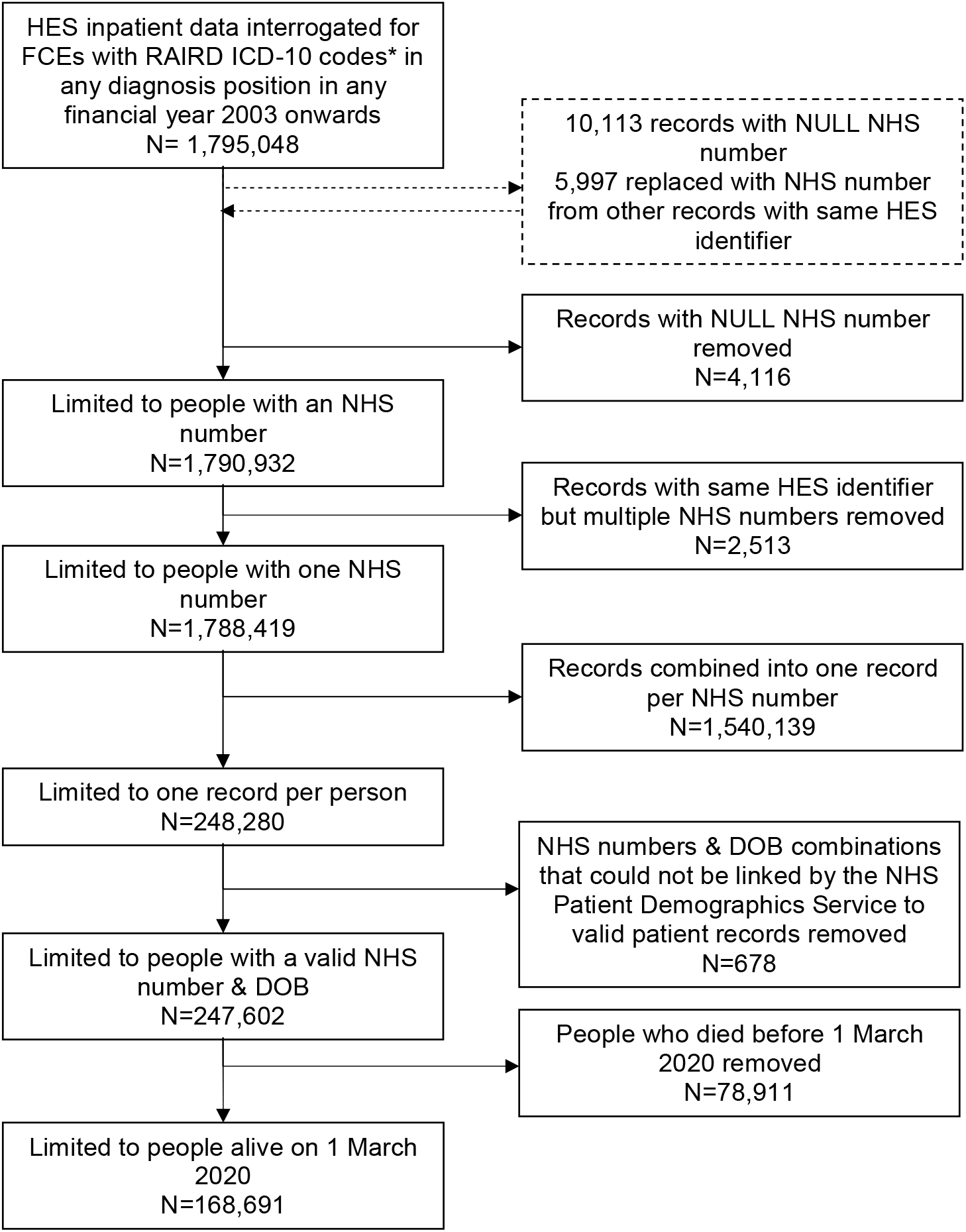
Flowchart of identification of the cohort of people with RAIRD *RAIRD ICD-10 codes include: M313, M317, M301, M314, I776, M352, M315, M316, M321, M330, M332, M331, M339, M340, M341, M348, M349, M083, M084, M082, M080, M300, M308, J991, N085, N164, M328, M329, M609, G724, M608, M089

We accessed publicly available data from the ONS statistical bulletin on “Deaths involving COVID-19, UK: deaths occurring between 1 March and 30 April 2020” to compare our crude, age-standardised and age-specific mortality rates to the whole population of England during the same time period(7).

This study received a favourable opinion from the Camden and Kings Cross Research Ethics Committee, study reference 20/HRA/2076, on 18 June 2020. Data were accessed and processed under section 251 permission granted to NCARDRS CAG 10-02(d)/2015). For quality assurance the data extraction and analysis were re-conducted by an independent analyst from the National Cancer Registration and Analysis Service (NCRAS).

Patient and public involvement: This project was discussed with and the aims supported by the Rare Autoimmune Rheumatic Disease Alliance of patient charities(8). A lay summary is available as an online supplement, and the results are being discussed with patients and representatives of the RAIRDA charities and disseminated to patients via their networks.

## Results

We identified 168,691 people with RAIRD diagnoses in HES who were alive on 1 March 2020 (figure 1). Validation of diagnoses was completed in 743 sets of case notes. These included at least 40 patients with each diagnosis from at least 2 hospital Trusts (except where the code occurred so infrequently that fewer than 20 cases were coded in each Trust). The results are shown in supplementary data Table 1. Overall, when weighted for the actual occurrence of the codes in the study population, the positive predictive value of our case ascertainment strategy was 84.7%. The characteristics of the cohort are shown in table 1.

**Table 1.** Characteristics of the cohort of people with RAIRD alive on 1 March 2020

| Characteristics of the RAIRD cohort | N (%) |
| --- | --- |
| Total number | 168,691 (100%) |
| Males* | 50,311 (29.8%) |
| Females* | 118,379 (70.2%) |
| Age on 1 March 2020 – Median (IQR) | 61.7 (41.5-75.4) |
| Died during March & April 2020 | 1,815 (1.1%) |
| *Most recent HES diagnosis* |  |
| Giant cell arteritis | 37,214 (22.1%) |
| Systemic lupus erythematosus | 36,419 (21.6%) |
| Juvenile inflammatory arthritis | 21,114 (12.5%) |
| Arteritis unspecified | 20,326 (12.1%) |
| Polymyositis | 17,041 (10.1%) |
| Scleroderma | 10,582 (6.3%) |
| Behcet's disease | 4,771 (2.8%) |
| Glomerular disorder in systemic connective tissue disease | 4,739 (2.8%) |
| Granulomatosis with polyangiitis | 3,997 (2.4%) |
| Respiratory disorder in other diffuse connective tissue disease | 2,825 (1.7%) |
| Dermatomyositis | 2,395 (1.4%) |
| Polyarteritis nodosa | 2,006 (1.2%) |
| Eosinophilic granulomatosis with polyangiitis | 1,980 (1.2%) |
| Renal tubulo-interstitial disorder in connective tissue disease | 1,110 (0.7%) |
| Microscopic polyangiitis | 918 (0.5%) |
| Takayasu's disease | 773 (0.5%) |
| Juvenile myositis | 480 (0.3%) |

\*1 patient did not have their sex recorded, and was excluded from male/female results.

During March and April 2020 1,815 (1.1%) of people with a RAIRD died of any cause. Crude and age-standardised mortality rates are shown in table 2. The crude mortality rate was 6481.4 (95% CI 6190.0-6786.5) per 100,000 person-years, and the age-standardised mortality rate (ASMR) was 3669.3 (3500.4-3838.1) per 100,000 person-years. The mortality rate in March and April 2020 was higher than during March and April over the previous 5 years (Mean ASMR for March and April 2015-2019 was 2554.2). The ratio of the ASMR in March and April 2020 compared to the mean ASMR in March and April 2015-2019 was 1.44 (95% CI 1.42-1.45).

**Table 2:** Deaths and mortality rates in England during March and April 2015-2020 among people with RAIRD compared to the general population

| Year | Number of deaths | Number of people | Person years | Crude mortality rate per 100,000 person-years | RAIRD Age standardised mortality rate | England age standardised mortality rate |
| --- | --- | --- | --- | --- | --- | --- |
| 2020 | 1,815 | 168,691 | 28,003 | 6481.4 (6190.0-6786.5) | 3669.3 (3500.4-3838.1) | 1361.1 (1353.6-1368.7) |
| 2019 | 1,236 | 159,378 | 26,257 | 4707.3 (4452.1-4977.2) | 2256.9 (2131.1-2382.7) | 5-year UK average<br>983.3 (980.3-986.3) |
| 2018 | 1,304 | 148,035 | 24,365 | 5351.9 (5069.2-5650.4) | 2618.8 (2476.7-2760.9) |  |
| 2017 | 1,101 | 137,628 | 22,677 | 4855.1 (4576.6-5150.5) | 2364.7 (2225.0-2504.4) |  |
| 2016 | 1,116 | 127,608 | 20,981 | 5319.2 (5016.1-5640.6) | 2953.0 (2779.7-3126.2) |  |
| 2015 | 1,004 | 117,450 | 19,300 | 5202.1 (4890.1-5534.0) | 2577.5 (2418.1-2736.9) |  |

**Table 3:** Sex-specific deaths and mortality rates in England during March and April 2015-2020 among people with RAIRD compared to the general population

| Sex | Number of deaths | Number of people | Person years | Crude mortality rate per 100,000 person-years | RAIRD Age standardised mortality rate | England age-sex standardised mortality rate |
| --- | --- | --- | --- | --- | --- | --- |
| Males | 675 | 50,311 | 8340.7 | 8092.8 (7504.8-8727.0) | 4025.4 (3721.7-4329.0) | 1626.7 (1613.8-1639.6) |
| Females | 1140 | 118,379 | 19,662.4 | 5797.9 (5470.9-6144.4) | 4242.1 (3995.9-4488.4) | 1150.7 (1141.5-1159.9) |

In the whole population of England, the ASMR reported by the ONS for the UK during March and April 2020 was 1361.1 (1353.6-1368.7) per 100,000 people, compared to 3669.3 (3500.4-3838.1) per 100,000 person-years among people with RAIRD. The 5-year average for March and April 2015-2019 in England was 983.3 (980.3-986.3) per 100,000. The ratio of the 2020 value to the 5-year average was 1.38. The ratio of the ASMR in 2020 to the average of the previous 5 years was a little higher in people with RAIRD (1.44; 95% CI 1.42-1.45) compared to the whole population (1.38) and 2.7 times higher than the ASMR for the general population of England during March and April 2020, compared to 2.6 times higher on average over the previous 5 years.

### Sex-specific mortality rates

The ASMR for males was 4025.4 (3721.7-4329.0) per 100,000 person years, and for females was similar at 4242.1 (3995.9-4488.4). This is in contrast to the general population where the ONS report males had a higher overall ASMR of 1,626.7 (1613.8-1639.6) deaths per 100,000 population compared with 1150.7 (1141.5-1159.9) for females.

Methodological note: Applying the European standard population which is not disaggregated by sex, (meaning it assumes equal numbers of males and females, and identical distributions by age for males and for females) means the age-sex standardised mortality rates are higher than the age-standardised mortality rate for this distribution of data.

### Age-specific mortality rates

Age-specific mortality rates during March and April 2020 in people with RAIRD compared to the whole population of the UK (data for England were not available) are shown in Figure 2 and Table 4. The rates of death among people with RAIRD were higher at each age-group than in the general population, and higher during COVID-19 than in 2019. The 20-29 years age-group was the youngest age-group we could report on due to small numbers in the younger groups. The diagnoses of the 65 people with RAIRD who died aged <50 years during March and April 2020 during COVID-19 are shown in supplementary data table 2.

**Table 4:**
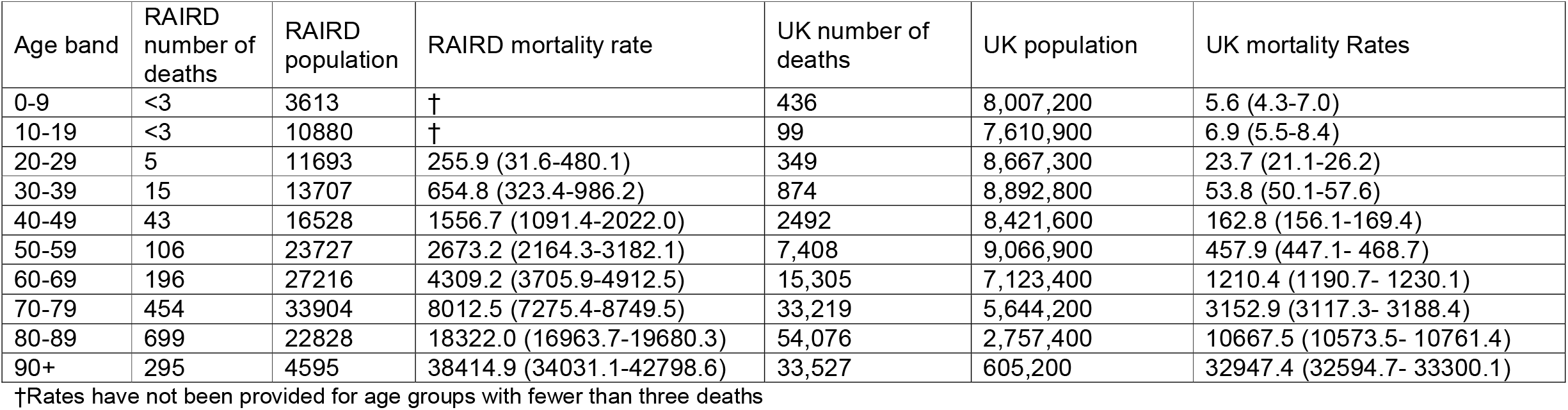
Age-specific mortality rates in people with RAIRD compared to the whole UK population

**Figure 2:**
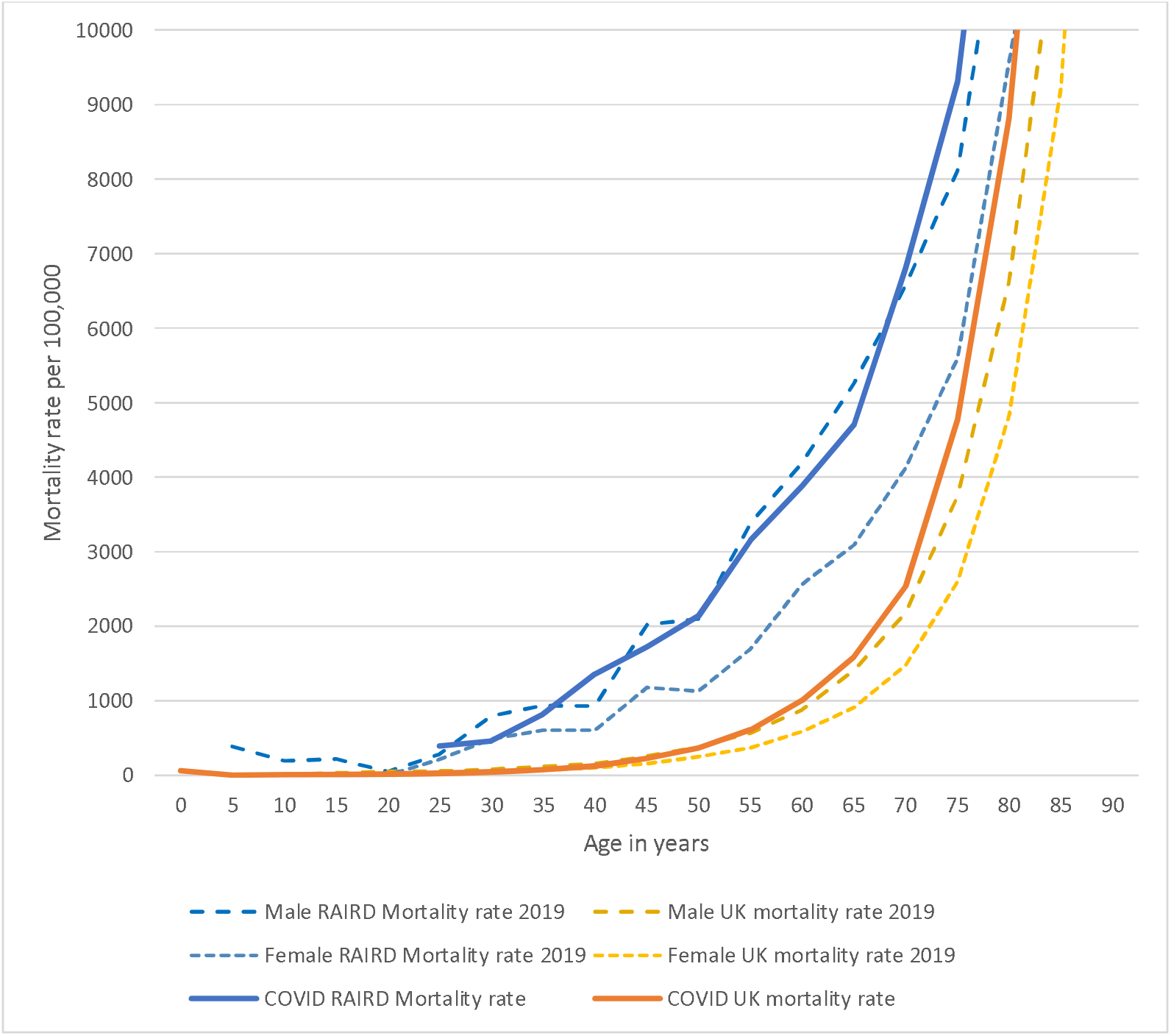
Age-specific mortality rates in people with RAIRD compared to the whole UK population during March/April 2020 and in 2019.

## Discussion

We provide the first evidence of the risk of death during COVID-19 in people with rare autoimmune rheumatic conditions compared to the risk not during COVID-19, as well as compared to the general population. The increased risk of death compared to the pre-COVID risk among people with RAIRD was slightly higher than in the general population. The risk was similar in males and females, in contrast to the risk in the general population where it is higher in males. When comparing risks of death during COVID-19 to pre-COVID-19, people with RAIRD had an increased risk of death from age 35 upwards, whereas in the general population the increased risk occurred from around age 55 upwards. Because the protective effect of being female is not seen in RAIRD, the group at the largest increase of risk compared to their pre-COVID-19 risk were women aged 35 upwards. The absolute risk of all-cause death for someone aged 20-29 with RAIRD was similar to someone in the general population aged >20 years older, and 40-49 years with RAIRD similar to someone in the general population 20 years older, and someone aged 60-69 with RAIRD was similar to someone in the general population aged >10 years older.

One key strength of this study is our ability to use a validated method of population-based case ascertainment in the whole English population of 55 million. Rare diseases are difficult to study, because not many people are affected by each disease(10). The best way to study risk in rare diseases is in whole population-based studies, enabled now in England by the unique capability of the National Congenital Anomaly and Rare Disease Registration Service within Public Health England. We ascertained cases from HES, which has been shown to have high coding accuracy for diagnostic ICD codes(11). Our validation work confirms this methodology is robust, with PPV of 85%. Although some of our cases (about 15%) may not therefore have a rare disease, as a result they are likely to be at lower risk of poor outcome. This may attenuate any effect we detected, so our results should be seen as a lower limit/underestimate. Other weaknesses are that by identifying cases using in-patient data we may miss patients only treated as outpatients, which are not routinely coded in HES. We have included primary and co-morbid diagnoses to maximise sensitivity, and prevalence estimates based on our findings for AAV, systemic lupus erythematosus and scleroderma, are similar to reported population estimates(12–14).

Recently the European League Against Rheumatism (EULAR) reported “no evidence that people with rheumatic musculoskeletal disorders face more risk of contracting SARS-CoV-2 than individuals without rheumatic musculoskeletal disorders, nor that they have a worse prognosis when they contract it.” (15) No evidence was presented on the risk to people with RAIRD compared to the general population. Our study provides the first evidence of this risk, and suggests that people with rare autoimmune rheumatic diseases are at higher risk of death during COVID-19 than the general population. The recent OPENSAFELY study, reported on the risk of in-hospital death due to COVID-19 among >17 million community based adults(16). The analysis included “common autoimmune diseases” reporting the combined risk for people with rheumatoid arthritis, lupus or psoriasis. However, because it does not have whole population coverage, even the large OPENSAFELY study does not have a sufficiently large cohort to study rare diseases. The adjusted hazard ratio for common autoimmune diseases was 1.23 (95% CI 1.12-1.35) compared to our overall ASMR for RAIRD which was 2.7 times higher than the ASMR for the general population of England. OPENSAFELY report in hospital deaths due to COVID-19, whereas we report all-cause deaths, and OPENSAFELY included a wider group of people with autoimmune diseases, which are likely to confer a lower risk than the rare autoimmune rheumatic diseases.

There are clinical and policy implications from this research. People with RAIRD have an excess risk of death during COVID 19 which starts at a younger age than in the general population, and is particularly prominent in young females. It is possible that this increased risk relates to both specific COVID infection risks but also could be due to the effects of change in consulting behaviour, and disruption to NHS services for rare diseases. For newly diagnosed patients their long “diagnostic odyssey” is likely to have been prolonged even further by closure of departments to routine referrals and they may be more unwell at diagnosis. Barriers to ongoing care because of strict social distancing policies might also be particularly relevant, as this population has a high prevalence of shielding (17,18). This work supports the prioritisation of RAIRD highlighted in NHS England guidance for primary and secondary care, principles that need to continue during service restoration to reduce risk of avoidable harm to this group (19,20).

There are implications for future risk assessments. At present COVID-age, the most frequently used workplace risk assessment tool for COVID and does not include rare disease risks, and so would underestimate the risks particularly to young and/or female people with RAIRD(21). The planned COVID-19 risk prediction model commissioned by Office of the Chief Medical Officer for England to NERVTAG has published their protocol which includes both common and rare autoimmune rheumatic diseases grouped together with non-immune diseases such as Ehlers-Danlos syndrome which will underestimate the risk in RAIRD. In addition this group are often treated with immunosuppression or high cost biologic drugs prescribed or administered in hospital which will not be available in their datasets. The protocol also excludes a large group of young people with the RAIRD diagnoses of juvenile idiopathic arthritis and Takayasu’s arteritis; and other vasculitides such as granulomatosis with polyangiitis and eosinophilic granulomatosis with polyangiitis(22).

These results also highlight the unique capability of the National Congenital Anomaly and Rare Disease Registration Service to inform population level strategies to support and protect the health of people with rare diseases (23,24). Current risk stratifications broadly assume that young people are not at increased risk of poor outcome from COVID-19 and do not need to “shield”. Our initial results challenge this and also have implications for prioritisation of vaccination in at-risk groups. (25,26).

Further research is urgently needed on the causes of death to understand the how many people are dying of COVID-19 and how many due to other causes, as well as the effect of ethnicity, immunosuppression and steroid usage on risks for people with RAIRD.

## Conclusion

The excess risk of all-cause death during COVID-19 occurs at a younger age among people with RAIRD than among the general population and particularly affects females. We urgently need to quantify how much risk is due to COVID-19 infection and how much due to disruption to healthcare services to inform better shielding advice, NHS service priorities and vaccine priorities for people with rare diseases.

## Supporting information

lay summary

## Data Availability

NCARDRS data are available to all who have a legal basis to access them. Further information is available at by application to Public Health England Office for data release (Available from www.gov.uk/government/publications/accessing-public-health-england-data/about-the-phe-odr-and-accessing-data [cited 2020 Oct 9])

## Acknowledgements

We would like to thank Chetan Mukhtyar, Reem Al-Jayyousi, Bridget Griffiths, Richard Watts, Mithun Chakravorty, Julie Battista, Robin Glover, Matthew Bell and Kay Randall for their help confirming diagnoses in hospital medical notes.

This work uses data that has been provided by patients, the NHS and other health care organisations as part of patient care and support. The data is collated, maintained and quality assured by the National Congenital Anomaly and Rare Disease Registration Service, which is part of Public Health England (PHE).**Transparency declaration**: the final author (the manuscript’s guarantor) affirms that the manuscript is an honest, accurate, and transparent account of the study being reported; that no important aspects of the study have been omitted.

## Data availability statement

NCARDRS data are available to all who have a legal basis to access them. Further information is available by application to Public Health England’s Office for data release(9).

---

**Supplementary Table1:** Results of validating the diagnoses of a sample of cases ascertained, reporting the positive predictive value (PPV) and weighting results by the frequency of codes in the study cohort.

| **RAIRD** | **ICD-10 codes** | **No. of cases validated in notes** | **No. of confirmed cases** | **PPV** | **Weighting of diagnosis in our cohort** | **Contribution to weighted PPV** |
| --- | --- | --- | --- | --- | --- | --- |
| **Giant cell arteritis** | M315, M316 | 65 | 53 | 81.5% | 0.2206 | 18.0% |
| **Systemic Lupus Erythematosus** | M321, M328, M329 | 37 | 31 | 83.8% | 0.2159 | 18.1% |
| **Juvenile Idiopathic Arthritis** | M080, M082, M083, M084, M089 | 40 | 40 | 100.0% | 0.1252 | 12.5% |
| **Arteritis unspecified** | I776 | 38 | 25 | 65.8% | 0.1205 | 7.9% |
| **Polymyositis** | M332, M608, M609, G724 | 38 | 33 | 86.8% | 0.101 | 8.8% |
| **Scleroderma** | M340, M341, M348, M349 | 38 | 33 | 86.8% | 0.0627 | 5.4% |
| **ANCA-Associated Vasculitis** | M301, M313, M317 | 220 | 189 | 85.9% | 0.0408 | 3.5% |
| **Glomerular disorder in connective tissue disease** | N085 | 36 | 32 | 88.9% | 0.0281 | 2.5% |
| **Behcet's disease** | M352 | 36 | 26 | 72.2% | 0.0283 | 2.0% |
| **Respiratory disorder in other diffuse connective tissue disorder** | J991 | 38 | 34 | 89.5% | 0.0167 | 1.5% |
| **Dermatomyositis** | M331, M339 | 40 | 38 | 95.0% | 0.0142 | 1.3% |
| **Polyarteritis Nodosa** | M300, M308 | 32 | 29 | 90.6% | 0.019 | 1.7% |
| **Renal tubulo-interstitial disorder in systemic connective tissue disorder** | N164 | 7 | 7 | 100.0% | 0.0066 | 0.7% |
| **Takayasu arteritis** | M314 | 63 | 50 | 79.4% | 0.0046 | 0.4% |
| **Juvenile Dermatomyositis** | M330 | 15 | 15 | 100.0% | 0.0028 | 0.3% |
| **All RAIRDS** | All | 743 | 635 | **85.5%**<br>**(95% CI 82.7-87.9)** | **1** | **84.7%** |

**Supplementary Table 2:**
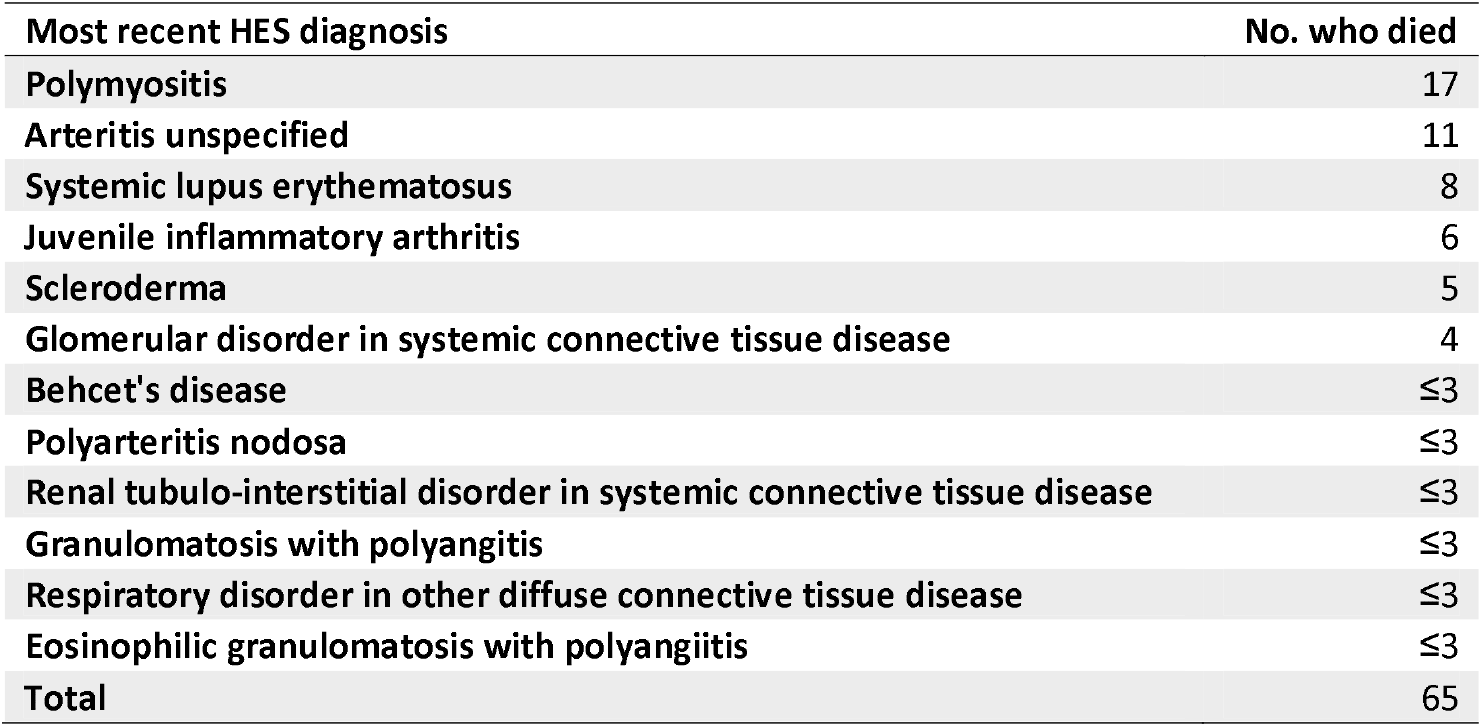
Diagnosis of the people with RAIRD who died under the age of 50 years during March and April 2020 during COVID-19

| **Most recent HES diagnosis** | **No. who died** |
| --- | --- |
| Polymyositis | 17 |
| Arteritis unspecified | 11 |
| Systemic lupus erythematosus | 8 |
| Juvenile inflammatory arthritis | 6 |
| Scleroderma | 5 |
| Glomerular disorder in systemic connective tissue disease | 4 |
| Behcet's disease | ≤3 |
| Polyarteritis nodosa | ≤3 |
| Renal tubulo-interstitial disorder in systemic connective tissue disease | ≤3 |
| Granulomatosis with polyangitis | ≤3 |
| Respiratory disorder in other diffuse connective tissue disease | ≤3 |
| Eosinophilic granulomatosis with polyangiitis | ≤3 |
| **Total** | **65** |

