## Supplementary material for "Risk of death among people with rare autoimmune diseases compared to the general population in England during the 2020 COVID-19 pandemic": lay summary

**Lay summary – How many people with rare autoimmune rheumatic diseases died during the 1st wave of the COVID-19 pandemic compared to the general population?**

We are a team of doctors and researchers from the RECORDER project (Registration of Complex Rare Diseases Exemplars in Rheumatology). This is a joint project between the University of Nottingham and the National Disease Registration Service at Public Health England. We looked at whether people with rare autoimmune rheumatic diseases, such as SLE (also known as lupus), scleroderma, vasculitis, juvenile idiopathic arthritis, myositis and Behcet's disease were affected more seriously during the COVID-19 pandemic than the general population, and whether people with these conditions had an increased risk of dying during this time.

People with rare diseases often have poorer health outcomes than people with more common diseases. A recent large UK study by OPENSAFELY found that people with common autoimmune diseases, such as rheumatoid arthritis or psoriasis, had a little increased risk of death due to COVID-19. However, not much is known about the risks to people with rare autoimmune diseases during COVID-19.

**Our findings:**

We looked at the electronic health records of nearly 170,000 people in England with rare autoimmune rheumatic diseases. During March and April 2020, the first two months of the COVID-19 pandemic in England, we found that 1815 (1.1%) people with these diseases died. Similar to other studies we found that the risk of death during COVID-19 increased more steeply with increasing age.

Our main findings were:

- When compared to before COVID-19, the risk of dying during COVID-19 for people with a rare autoimmune rheumatic disease increased from age 35 years onwards, whereas in the general population it increased from the age of 55 onwards.
- When adjusted for age, the risk of dying during COVID-19 was similar for men and women with a rare autoimmune rheumatic disease, whereas before COVID-19 women with rare autoimmune rheumatic diseases had a lower risk of death. This means women had a greater increase in their risk of death during COVID-19 compared to men.
- For people of working age with rare autoimmune rheumatic diseases, the risk of dying during the COVID-19 pandemic was similar to that of someone 20 years older in the general population.

**Future work:**

- We have not yet looked into the reasons why people have died. We will look into whether this was because of COVID-19, their rare autoimmune rheumatic disease, or another illness.
- This work does not show the effect of shielding, and whether things may have been worse without shielding. To answer this question we will look at rates of COVID-19 infection and the reasons why people died in people with rare autoimmune rheumatic diseases compared to the general population.
- The work will help to inform plans for shielding, how to deliver healthcare and prioritise services to keep open during a second wave of COVID-19. It will also inform future priorities for COVID-19 vaccination if and when a vaccine becomes available.

This is a summary of: ***Risk of death during the 2020 UK COVID-19 epidemic among people with rare autoimmune diseases compared to the general population. A whole-population study in England, using data from the National Disease Registration Service and the Registration of Complex Rare Diseases - Exemplars in Rheumatology (RECORDER) project*.** Emily J Peach, Megan Rutter, Peter C Lanyon, Matthew J Grainge, Richard Hubbard, Jeanette Aston, Mary Bythell, Sarah Stevens, Fiona A Pearce. **doi:** https://doi.org/10.1101/2020.10.09.20210237
